# A Cross-Sectional Study on Medication Errors in Home Health Care: The Role of Caregiver Type at a Military Hospital in Riyadh, Saudi Arabia

**DOI:** 10.64898/2026.03.10.26348036

**Authors:** Daniela Costa, Husam Ardah, Amjad Searya

## Abstract

**Background:** Medication safety in the home setting is becoming an increasingly important issue, particularly for older adults managing chronic illnesses. In Saudi Arabia, specifically within the National Guard Health Affairs (NGHA) system, medications are dispensed free of charge; however, the absence of a unified national tracking system for medication distribution may lead to safety risks. The risks are related to the duplicated amount of storage and expired or duplicate medications. Additionally, the support of untrained family members as informal caregivers may delay the implementation of safe medication management practices at home.

**Methods:** This cross-sectional study was conducted at a military hospital under the NGHA in Riyadh, Saudi Arabia. A structured medication form was used to assess key aspects of medication safety, including type of medication, timing, dosage accuracy, storage conditions, and caregiver type. Data from 503 patients and 2,515 individual medications were analyzed. Descriptive statistics were used for the sample, and the chi-square test was applied to examine the association between caregiver type and medication errors. Statistical analysis was performed using SAS 9.4.

**Results:** Most caregivers were informal (60.04%), and the average patient age was 79.8 years. Among all medications, 13.2% had dosage errors, 2% were expired, and 14.6% were improperly stored. A significant number (22.5%) were not administered at the correct time. The most common drug categories were antihypertensives, gastrointestinal agents, and sedatives. Medication errors were significantly higher among patients cared for by informal caregivers (46.01%) compared to formal caregivers (19.10%) (p < 0.0001).

**Conclusion:** Medication errors at home are a worldwide concern. Our findings highlight the need for caregiver training to improve home medication management and some policy reforms, like a national medication tracking system. This aligns with the importance of ensuring patient safety at home and the relevance of addressing this concern in Saudi Arabia.

## Background

It’s a fact that globally, the population is becoming older, as observed in Saudi Arabia. As life expectancy increases, the prevalence of chronic diseases rises, leading to greater demand for medical care and medication. The increase in life expectancy and the decrease in mortality rates will strain the healthcare system worldwide (1). Saudi Arabia’s population is expected to grow by around 55 million by 2050, increasing the demand for healthcare services such as home health care assistance (2,3). These population growth rates are also directly related to aging and living expenses, which cause more chronic conditions. So, in this case, caregivers’ roles are crucial to providing daily assistance and a better quality of life. Shifting care from institutional settings to home environments has become more common due to medical advancements and longer life expectancies (4). Researchers have shown that home care can reduce healthcare costs by at least 30%. However, for home care to be effective, the comfort and safety of patients and caregivers must be ensured, and the diversity of caregivers should be considered (5,6).

In Western countries, the demand for home nursing care has risen due to the increasing number of elderly individuals with chronic illnesses who prefer to remain in their homes. Ensuring medication is safe in home care is a significant challenge. Most of the elderly population suffers from cognitive and physical limitations and depends on caregivers to manage their medications. Although medication safety has been a priority for years, it remains a persistent issue (7,8). Keeping patients safe at home can improve patient satisfaction and quality of life and reduce costs for the healthcare system (9,10).

One of the biggest challenges in home health care is medication safety. Medication is a critical issue in healthcare, particularly for older adults with chronic conditions who often rely on home care services. Polypharmacy, the regular use of five or more medications daily, is a substantial factor in medication-related issues and increases the risk of these errors (11). Medication errors can be described as unintentional failures in the drug treatment process. They can occur at any stage and directly threaten patient safety. These errors may be related to humans or the process, not the drug’s inefficacy (12). The World Health Organization (WHO) has identified improper medication practices, such as incorrect storage, administration, or prescription, as a significant issue in healthcare. Studies show that up to 30% of patients are exposed to potential medication errors, particularly during drug administration (13-16).

Despite global awareness, there remains limited understanding of the specific barriers and supports for safe medication management in home care environments, particularly within Saudi Arabia’s healthcare system. Improving medication safety must address these systemic issues, involve better caregiver education, and emphasize effective communication among all parties involved in patient care (17,18).

Despite advancements in healthcare, there remains a limited understanding of the supports and barriers to safe medication management, especially in home care environments where patients, family members, and healthcare providers interact (19). Moreover, the scenario of Saudi Arabia is remarkably different from that of most other countries. Most of all, medication is free of charge from government hospitals, which can lead to benefits for the patients and prevent things such as extra medication. Another main characteristic is the lack of software medication, meaning no cross-medication data exists, which can cause medication duplication or medication interaction. Also, the medical record number is not even one for all the healthcare services around the country, but it changes according to each hospital (20-22).

During the patients’ home visits, one of the nurses’ roles is to check all their medication and educate them to see if any mistakes were made. Our study aims to assess the most frequent medication errors in patients’ homes and see if there is any correlation between the errors and the type of caregiver (formal vs informal). Based on the results, we also strive to create strategies and programs to minimize those errors. Furthermore, the results are expected to provide a factual evidence base for measuring and improving a better input into patient safety and quality of life. The goal is to create awareness, protect patients from the adverse effects of medication errors by promoting patient safety, and explore strategies by nurses to offer better home support.

## 2. Methods

### 2.1. Study Design

This research is a cross-sectional study. A template was used to assess the medication errors at patients’ homes. The study was conducted within the HHC program of National Guard Health Affairs in King Abdulaziz Medical City in Riyadh, one of the largest hospitals in the central region of Saudi Arabia, which receives cases from all social and economic classes. This HHC offers diverse specialized care to patients in their homes. It provides nursing care, medical, nutrition, occupational therapy, dietitian, and social services. The aim addresses patients’ physical, emotional, and social needs.

The study was reviewed and approved by the Departmental Research Committee of the National Guard Health Affairs (NGHA). All participants were informed that their participation was entirely voluntary and that their responses would be kept strictly confidential and anonymous. Written informed consent was obtained from all participants before data collection, under ethical research standards.

### 2.2. Participants

This study included all patients enrolled in the Home Health Care (HHC) program at the military hospital who were available during home visits and agreed to participate. Patients are referred to the HHC program by the Most Responsible Physician (MRP), and those who meet the program’s eligibility criteria are enrolled, with their information recorded in the hospital’s electronic medical record system, “BestCare.” From the first nurse visit to the patient’s home, their entire period of care is continuously monitored through this system, allowing accurate tracking of their clinical and caregiving data.

The inclusion criteria for this study comprised patients aged 18 years or older, of Saudi nationality, who had at least one chronic disease, were taking five types of medications simultaneously, were dependent on assistance for daily living activities, and had been enrolled in the HHC program for a minimum of four weeks. Each patient was required to have a single designated caregiver, either formal (a trained professional) or informal (a family member or untrained individual), to ensure consistency in data collection related to caregiving factors. Both male and female patients were included.

### 2.3 Procedures

Data was collected from 503 out of 1007 patients who met the eligibility criteria for the year 2024. A total of 2,515 medication registration forms were completed for this sample. Patients who did not meet the inclusion criteria were excluded from the study. Each patient was included only once, and no repeated data were recorded. Patients with more than one medication error were also included in the analysis to capture the full scope of errors occurring in the home setting.

Data collection took place from January 2024 to December 2024 using a standardized template developed by the Home Health Care (HHC) team. This form was specifically designed to assess and document the types of medication errors occurring in patients’ homes.

We followed the following steps to collect patient information. First, we identified the patients who satisfied the established criteria. Then, during the HHC nurse routine visits, the patients and caregivers were informed in detail about the research study and its importance to improve better practice and patient safety. Those who agreed to participate were asked to sign an informed consent form. Upon obtaining consent, the nurses proceeded with data collection. We secure all patients’ and caregivers’ confidentiality, and all information is handled with utmost privacy to ensure their anonymity and data security. After the consent was obtained, during the HHC routine visits, the nurses reviewed all medications in the patient’s home using the medication template form. They recorded whether any medication errors existed and identified which type of error was found. Each patient/caregiver pair was assessed only once to avoid duplication in the dataset.

The template served as the primary tool for collecting essential data related to medication safety in the home setting. This structured form was developed by the Home Health Care (HHC) team to systematically assess key factors associated with medication administration and caregiver involvement.

The template was organized into seven key segments. The first section identified the participants’ characteristics, such as age and gender. The second section identified the category of each medication to classify the type of drug being used. The third section assessed the timing of medication administration, determining whether each medication was given at the correct time. The fourth section evaluated the dosage, documenting whether the patient received the correct dose, an overdose/an underdose, or a missing dose. The fifth section examined whether the medication was stored appropriately within the home environment, and the sixth section assessed whether the medication expired. The final section identified the caregiver type, distinguishing between formal (trained healthcare providers) and informal (family members or untrained individuals) caregivers.

This template enabled a consistent and comprehensive evaluation of medication-related practices at home, allowing for a deeper understanding of common medication errors. Its structured format supported efficient data collection and facilitated expressive analysis aimed at improving patient safety in the HHC setting.

### 2.4. Statistical Analysis

All categorical data obtained from the medication form were calculated and presented as frequencies and percentages. Continuous data was summarized using means, standard deviations, medians, and interquartile ranges, as appropriate. Inferential statistics, including the chi-square test, were used to compare variables and assess associations. A p-value of less than 0.05 was considered statistically significant. All data were entered and analyzed using SAS version 9.4 (SAS Institute Inc., Cary, NC, USA).

## 3. Results

### 3.1. Sociodemographic Characteristics of Study Participants

The study included 503 patients enrolled in the Home Health Care program (Table 1). The mean age of participants was 79.8 years (± 8.96), with a median age of 79.0 years and an interquartile range of 74.0 to 87.0 years. Participants’ ages ranged from 48 to 99 years. Regarding caregiver type, 39.96% (n = 201) of patients were cared for by formal caregivers, while 60.04% (n = 302) had informal caregivers. The gender distribution was nearly balanced, with females representing 52.7% (n = 265) and males 47.3% (n = 238) of the study population.

**Table 1.** Descriptive statistics of Study Participants.

| Variables | Category | Total |
| --- | --- | --- |
| AGE | n | 503 |
| AGE | Mean (SD) | 79.8 ± 8.96 |
| AGE | Median (Q1,Q3) | 79.0( 74.00 , 87.00) |
| AGE | Min , Max | 48.0 , 99.0 |
| CAREGIVER | Formal | 201 ( 39.96% ) |
| CAREGIVER | Informal | 302 ( 60.04% ) |
| GENDER | FEMALE | 265 ( 52.7% ) |
| GENDER | MALE | 238 ( 47.3% ) |

#### 3.1.2 Medication Characteristics

A total of 2,515 medication entries were analyzed (Table 2). Regarding dosage accuracy, 86.8% (n = 2,182) of medications were administered at the correct dosage. However, 5.3% (n = 134) represents missing dosage, and 7.9% (n = 199) involved overdosage or underdosage. About medication expiration, 2.0% (n = 51) of the medications were found to be expired, while the vast majority, 98.0% (n = 2,464), were within their valid expiration dates. Regarding storage conditions, 85.4% (n = 2,147) of medications were stored appropriately, whereas 14.6% (n = 368) were found to be stored improperly. Assessment of administration timing showed that 77.5% (n = 1,950) of medications were given at the correct time, while 22.5% (n = 565) were administered at incorrect times. Regarding the types of medications used, the most frequently reported classes included antihypertensives (15.71%, n = 395), gastrointestinal agents (12.56%, n = 316), cardiovascular drugs (11.69%, n = 294), anxiolytics or sedatives (10.22%, n = 257), and antifungals (9.54%, n = 240). Other common medication classes included antidepressants (8.39%, n = 211), antipsychotics (6.60%, n = 166), hormonal medications (6.24%, n = 157), oral hypoglycemics (6.24%, n = 157), respiratory drugs (5.92%, n = 149), analgesics (3.62%, n = 91), and antivirals (3.26%, n = 82)

**Table 2.** Descriptive statistics: Medication characteristics.

| Variables | Category | Total |
| --- | --- | --- |
| DOSAGE | CORRECT | 2182 ( 86.8% ) |
| DOSAGE | MISSING DOSAGE | 134 ( 5.3% ) |
| DOSAGE | OVERDOSAGE/UNDERDOSAGE | 199 ( 7.9% ) |
| EXPIRE_DATE | NO | 2464 ( 98.0% ) |
| EXPIRE_DATE | YES | 51 ( 2.0% ) |
| MEDICATION_STORED_PROPERLY | NO | 368 ( 14.6% ) |
| MEDICATION_STORED_PROPERLY | YES | 2147 ( 85.4% ) |
| TIME | NO | 565 ( 22.5% ) |
| TIME | YES | 1950 ( 77.5% ) |
| TYPE_OF_MEDICATION | Analgesics | 91 ( 3.62% ) |
| TYPE_OF_MEDICATION | Antidepressants | 211 ( 8.39% ) |
| TYPE_OF_MEDICATION | Antifungal | 240 ( 9.54% ) |
| TYPE_OF_MEDICATION | Antihypertensives | 395 ( 15.71% ) |
| TYPE_OF_MEDICATION | Antipsychotics | 166 ( 6.60% ) |
| TYPE_OF_MEDICATION | Antiviral | 82 ( 3.26% ) |
| TYPE_OF_MEDICATION | Anxiolytics / Sedatives | 257 ( 10.22% ) |
| TYPE_OF_MEDICATION | Gastrointestinal agents | 316 ( 12.56% ) |
| TYPE_OF_MEDICATION | Hormonal medication | 157 ( 6.24% ) |
| TYPE_OF_MEDICATION | Hypoglycemics oral | 157 ( 6.24% ) |
| TYPE_OF_MEDICATION | Respiratory drugs | 149 ( 5.92% ) |
| TYPE_OF_MEDICATION | Cardiovascular drugs | 294 ( 11.69% ) |

### 3.2 Association Between Caregiver Type and Medication Errors

From table 3, it’s possible to observe a total of 2,515 medication records analyzed, 35.31% (n = 888) were associated with at least one medication error, while 64.69% (n = 1,627) showed no errors. When stratified by caregiver type, 46.01% of the medications administered by informal caregivers were associated with errors (n = 697 out of 1,515), compared to only 19.10% of medications administered by formal caregivers (n = 191 out of 1,000). Contrariwise, 80.90% of medications managed by formal caregivers were free from errors (n = 809), whereas the error-free rate among informal caregivers was lower, at 53.99% (n = 818). The difference in medication error rates between formal and informal caregivers was statistically significant, with a p-value of <0.0001, indicating a strong association between caregiver type and the medication error occurrence. From Figure 1, it’s possible to see the most frequent common errors found at the patient’s home.

**Table 3.** Association between caregiver and medication error.

| Table of CAREGIVER by MEDICATION_ERROR |  |  |  |  |
| --- | --- | --- | --- | --- |
| CAREGIVER(CAREGIVER) | (MEDICATION_ERROR) |  |  |  |
| Frequency<br>Percent<br>Row Pct<br>Col Pct |  |  |  |  |
|  | yes | no | Total | p-value |
| Informal | 697<br>27.71<br>46.01<br>78.49 | 818<br>32.52<br>53.99<br>50.28 | 1515<br>60.24 | <.0001 |
| Formal | 191<br>7.59<br>19.10<br>21.51 | 809<br>32.17<br>80.90<br>49.72 | 1000<br>39.76 |  |
| Total | 888<br>35.31 | 1627<br>64.69 | 2515<br>100.00 |  |

**Figure 1.**
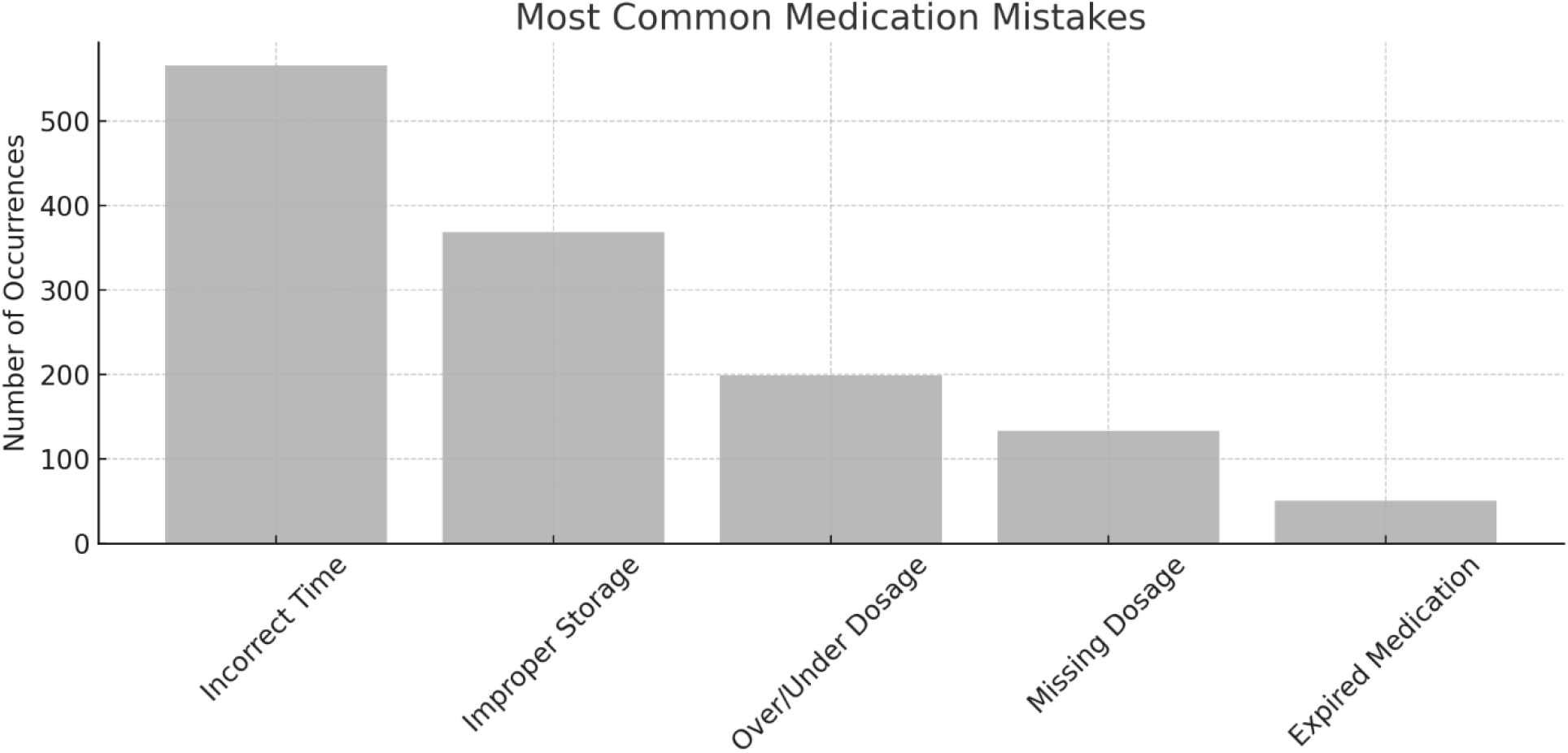
Description of the findings of medication errors.

## 4. Discussion

This study identified a considerable rate of medication errors among patients enrolled in the Home Health Care (HHC) program at a military hospital in Riyadh, Saudi Arabia. The most frequent types of errors were incorrect timing of administration, improper storage of medications, and dosing errors. Notably, a significant association was observed between the type of caregiver and the incidence of medication errors, with informal caregivers being associated with a higher error rate compared to formal caregivers (p < 0.0001).

Our findings are supported by previous research showing that informal caregivers are strongly associated with medication errors at patients’ homes (23). Comparing the types of caregivers (informal vs. formal), our findings show that formal caregivers are better prepared to understand medication schedules, dosages, and proper storage, which could be associated with their clinical training. On the other hand, informal caregivers, most often family members, who are not well trained or educated, demonstrated fewer skills to manage the patient’s medications, especially with elderly patients with multiple comorbidities and polypharmacy. This underscores the importance of training the caregivers, especially the informal ones, who need more health education and to be taught by expert health practitioners at their homes, particularly those with high-risk conditions.

Some cultural factors in Saudi Arabia could contribute to these medication errors. First, the NGHA institution’s system ensures that all medication provided is free of charge to all patients. This effectively ensures access to the patient’s needs, but it could negatively lead to duplicated medication storage at patients’ homes. Although not formally recorded in our tool, during data collection, it was frequently observed that patients had multiple packages of the same medication, including high-alert drugs such as narcotics. The lack of a centralized national health information system to track medication dispensing further exacerbates the risk of duplication and waste. Patients may request medications from different providers without an integrated oversight mechanism, resulting in excessive quantities of medication at home.

Expired medications in 2% reflect potential issues with inventory control and medication awareness among caregivers. In some cases, patients may continue to store and use medications that were no longer needed, simply because they were provided freely and were not discarded properly. This emphasizes the need for ongoing monitoring, education, and better communication between healthcare providers and caregivers regarding safe medication practices.

Our results are supported by prior literature, including studies from the Gulf region and broader international contexts, that highlight the risks associated with unregulated home medication management (24-26). Similar concerns about medication waste and the lack of tracking systems have been raised in another Saudi study (27), revealing that this is a national and generalized problem and not only in this specific institution.

Despite the valuable insights gained, this study has several limitations that must be acknowledged. First, the sample size was limited to 503 patients enrolled in a single military hospital’s Home Health Care (HHC) program. Although this population provides valuable insight into home care, the findings may not be generalizable to the broader Saudi population or other healthcare institutions. The small sample size also limits the statistical power to find deeper associations. It may not describe the full reality of medication errors occurring at patients’ homes.

Another limitation is the methodology based on a single point-in-time assessment during scheduled home care nurse visits. This design may have missed medication errors outside the observed period, such as intermittent or situational administration, storage, or timing mistakes. Moreover, while nurses recorded observed errors during their visits using a standardized form, there is a possibility of not correctly reporting or missing errors that caregivers did not immediately notice or disclose. Also, some observed practices by nurses during the patient’s home visits, such as the accumulation of duplicated medications, especially some high-alert drugs like narcotics, were not included officially in the data tool collection. Notwithstanding the importance of this matter, these observations that were not recorded were excluded from the quantitative analysis. Moreover, this specific context of the healthcare system in Saudi Arabia could not reflect the same thing that happens in other countries. This gap, in the free charge of medication and the enhancement of internal communications between healthcare institutions, leads to access to strong medication at patients’ homes. Probably, patients request medicines in more than one institution at the same time, especially narcotics, which causes not only duplicated drugs in the household but also increases the risk of harm and overdosing. Counting these limitations will be necessary for future research, including more sample diversity across other Saudi Arabian regions and different healthcare institutions. Longitudinal studies should be considered to provide a better picture and a comprehensive understanding of this medication error tendency. Additionally, future studies will greatly benefit as they assess the efficiency of target education interventions for the caregivers, such as training, administration, and medication management. Finally, collaboration with policymakers to support caregiver training and public awareness campaigns would be essential steps toward improving medication safety in the home setting.

## 4. Conclusion

This study highlights a significant prevalence of medication errors in the home healthcare setting within a military hospital in Riyadh, Saudi Arabia. Informal caregivers are associated with a higher error rate than formal caregivers. One of the most common medication errors was related to the time of administration, followed by the dosage inaccuracy and the improper storage. The findings underline the need for targeted interventions to reduce medication risks at home. It’s essential to develop standardized protocols to ensure proper medication management by training the caregivers, especially the informal ones. Moreover, the presence of expired medications and the observation of medication duplication at patients’ homes point to systemic gaps in medication tracking and disposal. These challenges are exacerbated by the absence of a unified national system to monitor medication dispensing and usage. This situation can result in oversupply or inappropriate access to high-alert medications such as narcotics. This is especially important in contexts where medications are dispensed free of charge, and the risk of accumulation and waste is high.

Finally, improving home-based medication management requires a multidisciplinary approach involving patients, caregivers, healthcare providers, and policymakers. Future efforts should focus on this research across different regions and care settings in Saudi Arabia to develop comprehensive strategies that ensure medication safety for all home healthcare recipients.

## Data Availability

All data produced in the present study are available upon reasonable request to the authors

## Consent for publication

Written informed consent was obtained from all participants before data collection, under ethical research standards.

## Availability of data and materials

The datasets used and/or analysed during the current study are available from the corresponding author on reasonable request.

## Contributions

Conceptualization D.C.; Methodology D.C.; Validation D.C, H.A, A.S.; Formal analysis D.C, H.A, A.S.; Investigation D.C.; Writing—Original draft preparation D.C.; Writing—review and editing D.C.; Visualization D.C.; Supervision D.C. All authors have read and agreed to the published version of the manuscript.

## Data sharing

The data presented in this study can be obtained by contacting the corresponding author if desired.

## Acknowledgments

We appreciate the support from all staff working in our home health care facility.

